# The Real-World Impact of Concussions on the Neuropsychological and Menstrual Health of Women

**DOI:** 10.64898/2026.08.21.26361020

**Authors:** Priya Ravi, Amanda Yad-El Ugboji, Grace Osborne, Maya Jokhadze, Beverly Oleka, Fizza Fatima, Celestin Niyomugabo, Meredith L Snook, Emma M Tinney, Goretti España-Irla, Kuo-Ting (Tim) Huang, Martina Anto-Ocrah

## Abstract

**Introduction:** While most concussion patients recover within one month, approximately 15 to 30% of patients experience prolonged symptoms. Women are disproportionately affected and report greater physical, cognitive, and mood-related symptoms than men. To date, much research on concussion and sex differences relies on quantitative measures, with limited exploration of longer-term female experiences. This study uses a mixed-methods approach to assess “real-world” symptom morbidity in terms of neurological, mental, and menstrual health approximately 2 years post-concussion, compared to non-head injured controls.

**Materials and Methods:** Eligible participants were i) assigned female at birth, ii) aged 18–45 years, iii) not using hormonal birth control, and iv) (for the concussion group) diagnosed within 7 days of injury. Participants were recruited (April 2023–September 2025) across University of Pittsburgh Medical Center sites, including the Concussion Clinic, Emergency Departments, Student Health Clinic, and Pitt + Me registry. Follow-up assessments occurred October–November 2025.

**Measures & Analyses:** Post-concussion symptom morbidity was assessed using the Rivermead Post Concussion Symptoms Questionnaire (RPQ), depression with the Patient Health Questionnaire-9 (PHQ-9), and anxiety with the Generalized Anxiety Disorder-7 (GAD-7). Menstrual health was assessed using study-specific measures developed by the research team with expertise in concussion and women’s health. Qualitative data were collected via open-ended responses on perceived life impacts and recurring themes were illustrated using word clouds generated from the frequency of reported responses.

**Results:** A total of 45 concussion patients and 29 controls were recruited; 11 concussion patients (mean age 30.4 ± 8.4 years) and 16 controls (mean age 31.3 ± 7.4 years) completed follow-up. Compared to controls at follow-up, concussion patients reported greater symptom burden 2(RPQ: 31.6 ± 13.5 vs 9.4 ± 9.8; p=0.0002; Hedge’s g=1.90), depression (PHQ-9: 9.5 ± 6.5 vs 2.3 ± 2.2; p=0.0005; Hedge’s g=1.60), and anxiety (GAD-7: 9.8 ± 6.8 vs 2.8 ± 3.0; p=0.0057; Hedge’s g=1.42).

Qualitative responses reinforced these findings: “No positive impacts. My relationship with my partner is downfall due to my lack of interest in intimacy and my lack of will. I’m drain[ed] every day of my life. I now use glasses as my vision is getting worse. My headaches are part of my everyday struggle. I can’t sleep at night as I have headaches or worries.” (C#9)

**Conclusions:** This mixed-methods study demonstrates significant long-term differences in symptom burden among women with concussions compared to controls. Findings highlight the importance of understanding real-world impacts to improve long-term care.

## Introduction/Background

Traumatic brain injuries (TBIs) affect an estimated 69 million people worldwide and are a leading cause of death and disability globally, posing a substantial public health burden.^1^ TBIs vary widely in severity and have been typically classified as mild, moderate, or severe based on clinical criteria, such as structural imaging findings, duration of loss or alteration of consciousness, post-traumatic amnesia, and Glasgow Coma Scale scores.^2^ In order to more accurately diagnose the types of TBI, this classification has begun to shift to a new multidimensional framework known as CBI-M that is now based on four categories: clinical information, biomarkers, imaging, and modifier features.^3^ Mild traumatic brain injury (mTBI), commonly referred to as *concussion*, is the most prevalent form of TBI, accounting for approximately 75% of all cases.^4^ The Mild Traumatic Brain Injury Committee defines mTBI as “a traumatically induced physiological disruption of brain function” that does not meet the criteria for moderate or severe TBI.^5^ Reported incidence rates among hospitalized patients range from 200 to 300 per 100,000 persons annually^6^; however, this likely underestimates the true burden, as many individuals do not seek medical care following injury.^7^ Underreporting may be due to limited symptom recognition and/or acknowledgement,^8^ a potential byproduct of gaps in follow-up care and concerns regarding financial or occupational consequences with missed work.^9,10^

Despite its classification as “mild,” concussions are frequently associated with a range of persistent symptoms, including headache, fatigue, mood disturbances, irritability, and cognitive impairment.^11^ Although many individuals recover within weeks, one in 4 patients experience prolonged or chronic symptoms that significantly impact daily functioning and quality of life; a condition characterized as persistent post-concussive symptoms (PPCS).^12^ Much of the literature on PPCS has historically aggregated men and women. However, emerging studies have identified the female sex as a predictor of PPCS,^13^ noting that compared with male counterparts, women consistently report higher post-concussion symptom burden, longer recovery times, and greater cognitive and neurosensory impairments in the months, at times years, following the initial injury.^14^ They are also more likely to experience changes in affect, including worsened depression, anxiety, and PTSD.^15,16^ Additionally, some studies have suggested that mild TBI can lead to abnormal menstrual patterns, potentially due to transient ischemic damage to the hypothalamus, infundibulum, and/or pituitary gland; acute hyperprolactinema; and activation of the hypothalamic-pituitary-adrenal axis owing to the stress response of an injury.^17,18^ A number of sex and gender-specific factors have been linked to both risk of concussion and recovery thereafter, including biological and anatomic factors such as hormonal differences and smaller neck size,^16^ social factors such as interpersonal violence and psychosocial stress,^19,20^ and behavioral factors such as innate differences in somatic perception, socialization, and willingness to disclose discomfort.^21^

Although there is recognition of these sex-based differences, much of the evidence base remains grounded in quantitative methods, relying heavily on standardized symptom inventories and self-report scales.^13^ However, addressing this gap requires not only separation of data by sex/gender but also a broader reconsideration of *how* these outcomes are measured. While quantitative approaches are essential for identifying statistical trends, they tend to constrain patient experiences into predefined categories and may fail to capture the complexity of recovery in everyday life.^22^ For example, symptom severity scores, although informative, do not necessarily reflect functional status, as they may not correspond to how individuals navigate daily activities, responsibilities, or social roles.^23^ Additionally, the emphasis on standardization with quantitative research often limits contextual information, excluding important social, psychological, and environmental factors that may shape patients’ recovery experiences.^24,25^ For women, this includes their menstruation experiences, which can influence both their physical and emotional wellbeing. Even with the emerging evidence on the impact of concussion on women’s menstrual health, there is limited research on their real-world experiences.^17^ Rather than functioning in opposition, qualitative and quantitative methods can be complementary, with qualitative research providing critical insight into the “how” of patients lived experiences, providing a more comprehensive understanding of post-concussion recovery.^24,26^ By centering patient narratives, qualitative approaches can capture dimensions of recovery that extend beyond numerical symptom burden, including identity and the lived realities of daily life following injury.^27^

The temporal scope of existing research further constrains understanding of women’s post-concussion experiences. Post-concussion recovery has been measured largely in the weeks to months following injury, reinforcing the notion that recovery is often reached within these timescapes.^28,29^ However, a subset of individuals has been shown to experience symptoms beyond this window, with evidence pointing to persistent cognitive, physical, and mental effects on quality of life.^30^ These longer-term recoveries remain insufficiently characterized, despite their relevance for women, who as previously mentioned, are more likely to experience prolonged symptom burden post-concussion.^14^ Examining experiences at least 1 year post-injury would allow for a more comprehensive understanding of recovery as a dynamic and ongoing process with changes that are not visible in early recovery windows.^31^

In this study, we use a mixed-methods approach to examine the long-term impact of concussion on women’s lived experiences in the real world. Women with a history of concussion were compared with non-head injured controls to specifically evaluate differences in:

1. post-concussive neurologic symptoms and mood-related disorders and ii) menstrual patterns, experiences, and changes in bleeding severity.

We hypothesized that the concussion group would report greater symptom burden, higher levels of psychological distress, and more frequent menstrual irregularities. By integrating statistical analysis with patient narratives, this study aims to identify key areas for improving clinical care, manage chronic symptoms (including those under menstrual health), and inform future research on sex-specific outcomes after concussion.

We acknowledge that although biological sex refers to the relatively stable biological characteristics associated with being male or female, gender encompasses the socially constructed roles, behaviors, expectations, and norms attributed to women, men, and people of various genders within a given society. For example, the capacity to bear children is determined by biology, whereas expectations surrounding childbearing, parenting responsibilities, and the social status associated with motherhood are shaped by gender.^32,33,34^ Consistent with this distinction, our manuscript is framed to explore gender—specifically how gendered roles, expectations, and social experiences influence the phenomenon under study; as well as sex-specific attributes such as menstruation.

## Methods

### Participants

#### Phase 1

Between April 2023 and September 2025, we recruited study participants from multiple sites at the University of Pittsburgh Medical Center (UPMC) including the Concussion Clinic, Emergency Departments, and the university’s Student Health Clinic. Additional participants were identified through Pitt + Me, a research registry of more than 300,000 individuals in the Pittsburgh region who have opted in to being contacted for future studies.

**Inclusion criteria:**

- Assigned female sex at birth,
- Aged ≥ 18 to ≤ 45 years
- Seeking care within ≤7 days after injury
- Not admitted as an inpatient after treatment
- Not currently pregnant/undergoing fertility treatments /using hormonal birth control.
- Additionally for non-head injured controls:

- No history of concussions within the last 6 months
- If extremity injured control (i.e. legs, arms), injury needed to be a) isolated extremity injury (i.e. legs, arms) b) with no signs of fracture on x-ray examination and no surgical intervention

**Exclusion criteria:**

- Unable to provide consent
- Has soft tissue injury such as ACL tears, post-ops, etc.

#### Phase 2

From October to November 2025, we followed up with the Phase 1 participants.

### Measures

#### Persistent Post-concussion Symptoms (PPCS)

We used the Rivermead Post-Concussion Questionnaire (RPQ)^13^ to evaluate PPCS. The RPQ is a 16-item self-report measure of the presence and severity of the 16 most commonly reported post-concussive symptoms observed in the literature. The scale compares any current symptoms to pre-injury symptom levels to account for potential symptom exacerbation subsequent to the head injury. Values for each of the 16 items are ranked on a five-point scale (0 = not experienced at all, 4 = severe problem). Scores on the RPQ range from 0 to 64, with higher scores suggestive of greater PCS burden. This scale has been endorsed for mTBI populations by the National Institutes of Health (NIH) division of National Institutes of Neurological Disorders and Stroke (NINDS), and used previously to assess outcomes in mTBI populations.

#### Depression

The Patient Health Questionnaire (PHQ-9) is a validated assessment of depression, with a numerical scoring system of 0–27.^35^ Higher scores indicate higher levels of depressive symptoms. Participants respond to how often they have been bothered by symptoms, from 0 (not at all) to 3 (nearly every day). Cutoffs are 0–4: Minimal depression, 5–9: Mild depression,10–14: Moderate depression, 15–19: Moderately severe depression, 20–27: Severe depression.

#### Anxiety

The GAD-7 is a validated questionnaire that assesses anxiety, with a score range of 0–21, with higher scores indicating higher levels of anxiety.^36^ Cutoffs are: 0–4: minimal anxiety, 5–9: mild anxiety, 10–14: moderate anxiety, 15–21: severe anxiety.

#### Menstruation experiences

Menstrual health measures were designed by the study’s reproductive epidemiologist (MAO).^37^ Questions asked about participants’ experiences with their menstrual flow, frequency, and pain in the last 3 months. The 3 month time frame was selected to standardize participants’ frame of reference for their responses.

#### Qualitative responses

We asked concussion participants to describe their lived experiences using open-ended text boxes. Specifically, we asked them to describe:

a. The positive and negative experiences with persisting concussion symptoms, if any
b. Their menstrual experiences compared to before their concussion, again citing any positive and negative impacts

#### Data Analyses

Quantitative data was analysed using Python version 3.11.9. We used descriptive statistics (proportions, means, medians, ranges, and standard deviations) to describe the study sample. Fisher’s exact test was used to compare categorical survey responses in bivariate analyses. Independent-samples t-tests were used to compare continuous variables when normally distributed, with Mann-Whitney U tests used otherwise. The statistical significance was set at p < 0.05. Qualitative responses were visualized using word clouds based on the frequency of reported themes, with word size scaled to the number of mentions. Word frequencies were computed using CountVectorizer (scikit-learn), and word clouds were generated using the wordcloud package. The study was approved by the University of Pittsburgh IRB (STUDY25060143).

## Results

In Phase 1 (baseline study enrollment), we recruited 45 concussion and 29 non-head injured controls for a total of 74 participants. Seventy two opted to be recontacted for future research and were recontacted to participate in Phase 2. Twenty seven (11 concussion and 16 non-head injured controls) participated in the follow-up study (Phase 2). As shown in Table 1, at the time of follow-up (Phase 2), the majority of the concussion group (63.6%) indicated that they were between 1 and 2 years post-injury. When asked about past concussion histories in Phase 2, 4 of the 16 controls were affirmative: 1 had sustained a concussion in the last 7-12 months, 2 had been injured 3-5 years ago, and 1 had their injury over 5 years ago. Of the concussion participants, 63.6% (n=7) with mental health histories at baseline enrollment (Phase 1) also participated in the follow-up Phase 2, compared to 37.5% (n=6) of controls (p=0.18). All of the controls in Phase 2 (n=16, 100%) and 91% of the concussion participants (n=10) had normal menstrual cycles (8-14 cycles/year) at baseline enrollment (Phase 1).

**Table 1:**
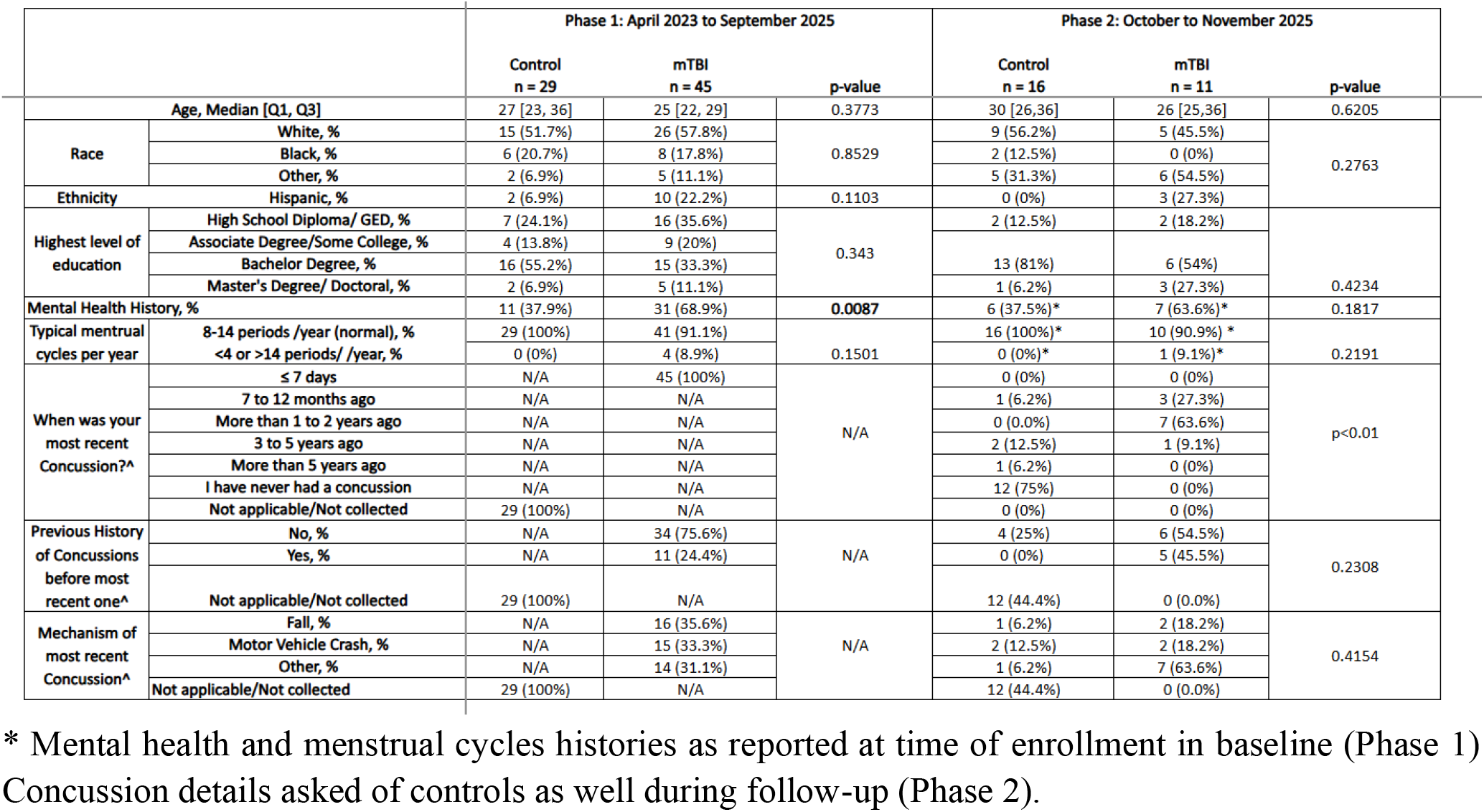
Demographic, mental health, menstrual health and injury-related attributes of control and concussion(mTBI) participants at baseline (Phase 1) and follow-up (Phase 2).

### Aim 1: Post-concussive neurologic symptoms and mood-related disorders

As shown in Table 2, the concussion group at follow up had a statistically higher burden of concussion symptoms, depression and anxiety than the non-head injured controls.

**Table 2:** Group Differences in PPCS, Depression, and Anxiety 1-2 years (on average) post-concussion.

| Measure | Concussion<br>(n=11) | Control (n=16)<br>(Mean +/-SD) | p-value | Hedge's g |
| --- | --- | --- | --- | --- |

|  | (Mean +/-SD) |  |  |  |
| --- | --- | --- | --- | --- |
| RPQ (PPCS symptoms) | 31.6 ± 13.5 | 9.4 ± 9.8 | 0.0002 | 1.90 |
| PHQ-9 (depression) | 9.5 ± 6.5 | 2.3 ± 2.2 | 0.0005 | 1.60 |
| GAD-7 (anxiety) | 9.8 ± 6.8 | 2.8 ± 3.0 | 0.0057 | 1.42 |

Figure 1 and Table 3 show the language used by participants to express their lived experiences with PPCS. Several comments included experiences with headaches/migraines, visual impairments, cognitive, and mental health impacts.

**Figure 1:**
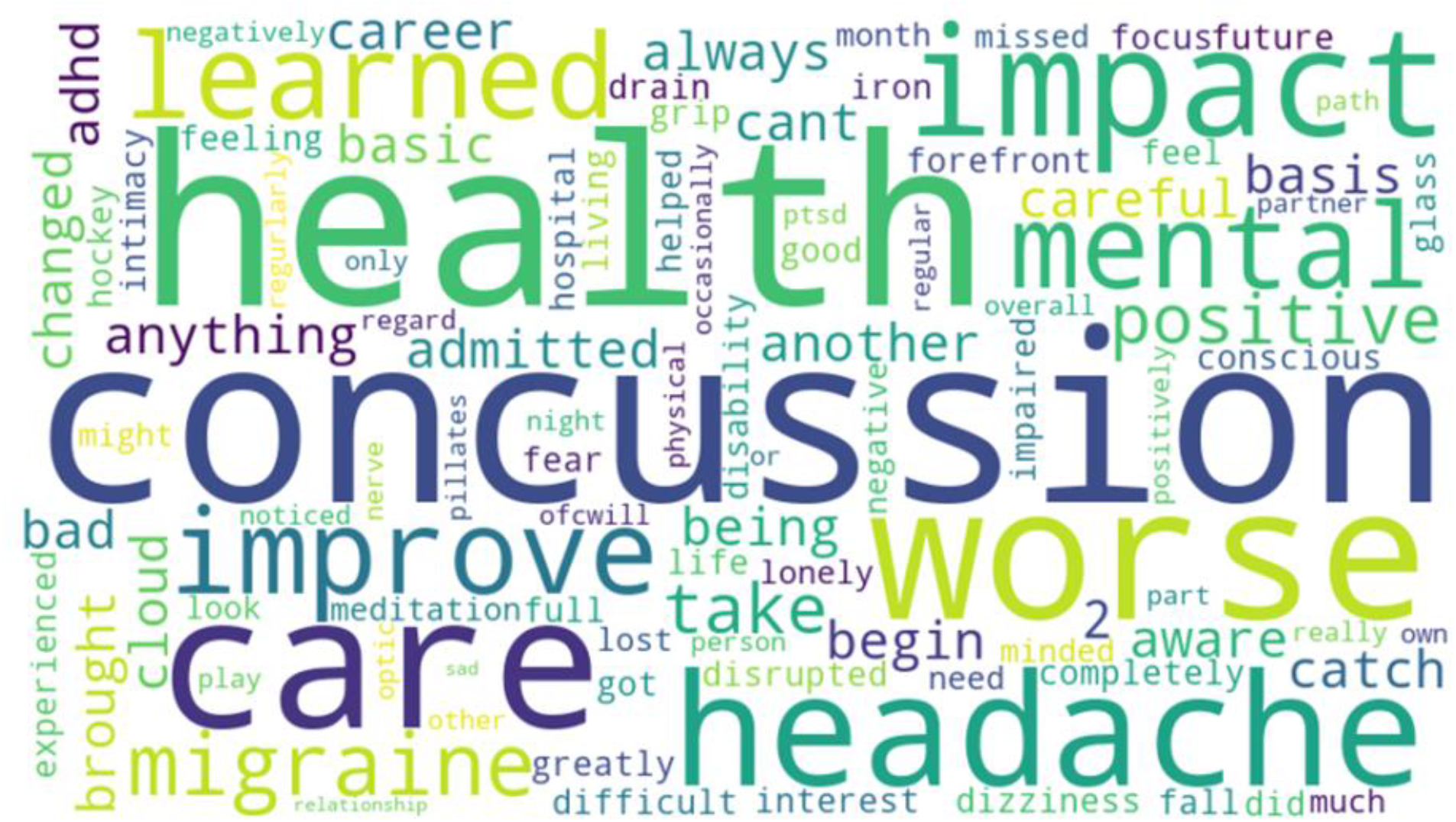
Word cloud visualization of emergent themes describing PPCS burden for concussion patients 1-2 years (on average) post-concussion. Weight and size correspond to the number of mentions (n=10).

**Table 3:**
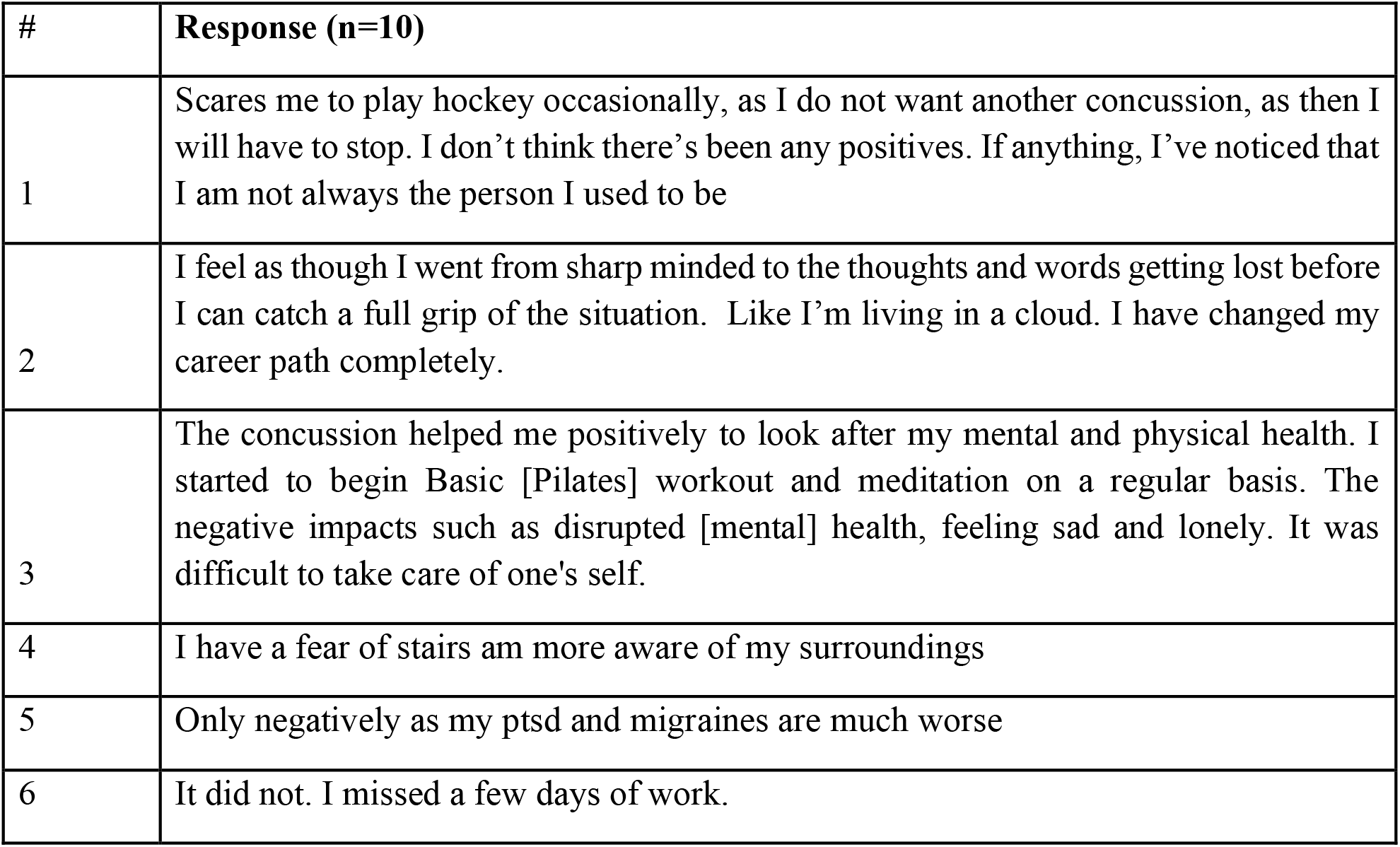
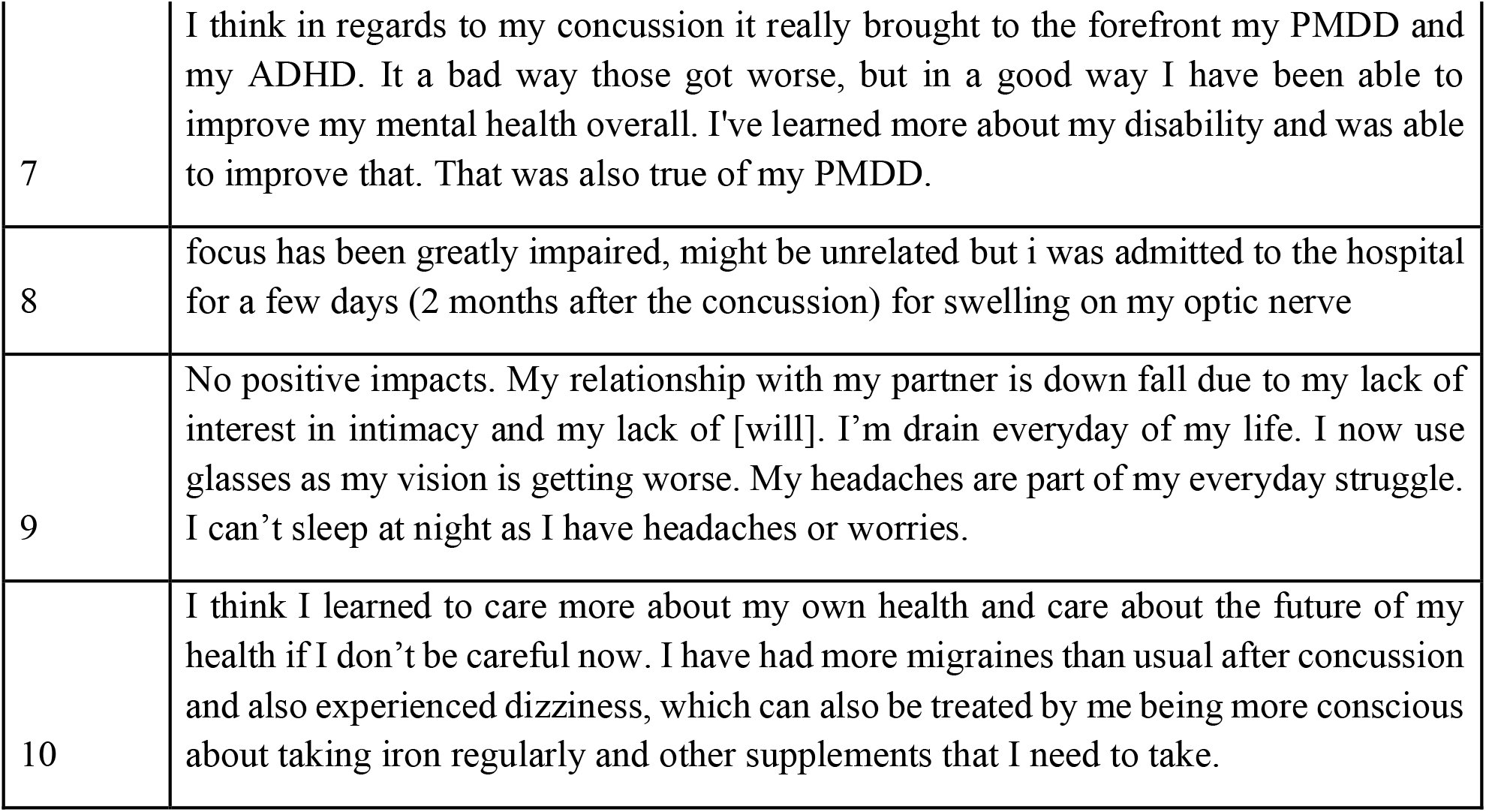
Language used by participants to express their lived experiences with PPCS 1-2 years (on average) post-concussion.

**Table 4:**
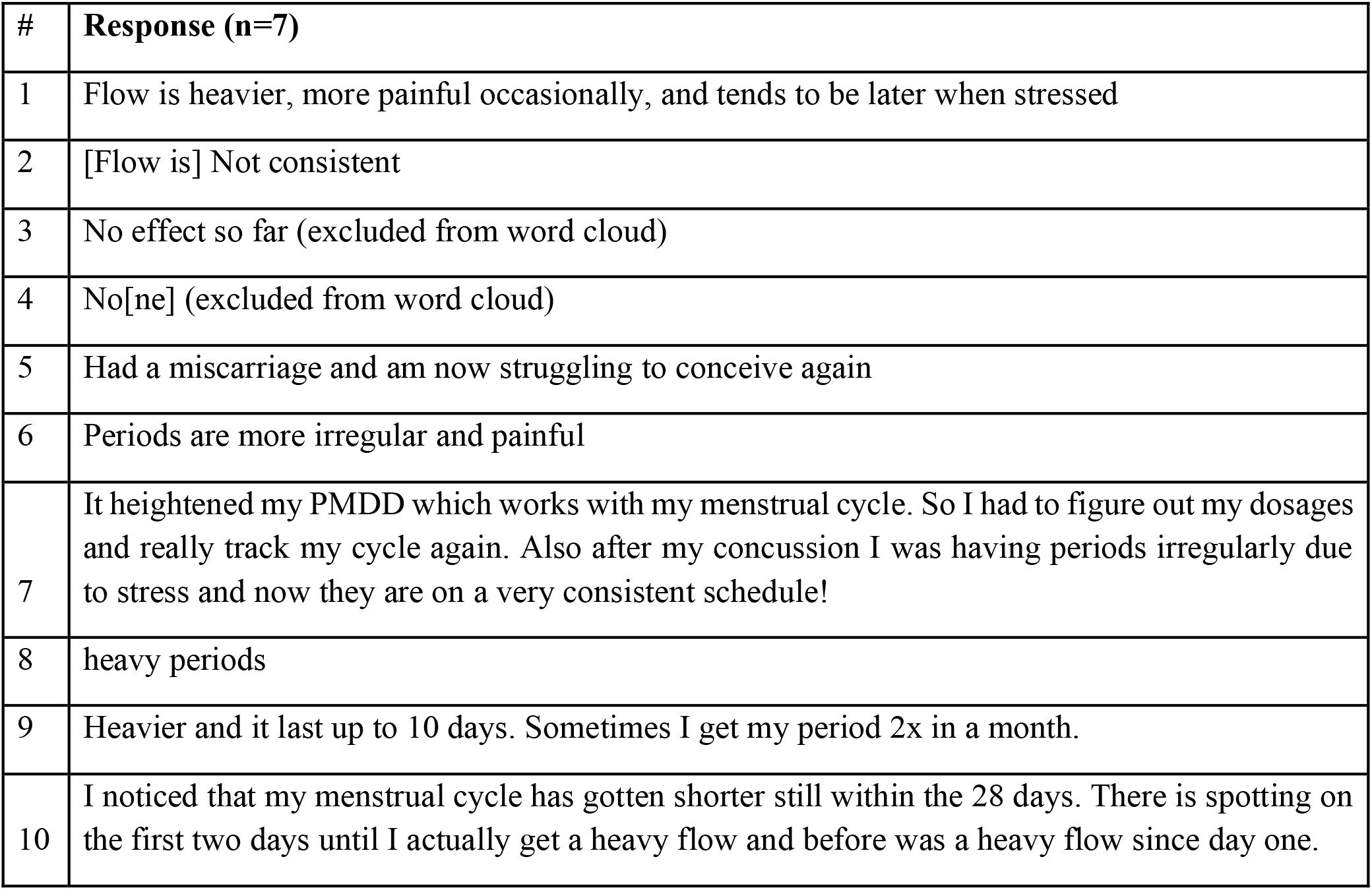
Language used by concussion patients to describe their menstruation experience 1-2 years (on average) post-concussion.

As a participant stated,

“Only negatively as my ptsd and migraines are much worse” (Response #5)

Another commented,

“focus has been greatly impaired, might be unrelated but i was admitted to the hospital for a few days (2 months after the concussion) for swelling on my optic nerve” (Response #8)

Another added,

“I feel as though I went from sharp minded to the thoughts and words getting lost before I can catch a full grip of the situation. Like I’m living in a cloud. “ (Response #2)

And finally,

“No positive impacts. My relationship with my partner is down fall due to my lack of interest in intimacy and my lack of will. I’m drain everyday of my life. I now use glasses as my vision is getting worse. My headaches are part of my everyday struggle. I can’t sleep at night as I have headaches or worries.” (Response #9)

Interestingly, a few reflected more positively

“The concussion helped me positively to look after my mental and physical health. I started to begin Basic Pilates workout and meditation on a regular basis” (Response #3)

“In a good way I have been able to improve my mental health overall. I’ve learned more about my disability and was able to improve that.” (Response #7)

Overall, participants reported more negative than positive impacts of PPCS.

The quantitative data from the RPQ supported these qualitative reports, with regards to severe, statistically significant differences in symptomatology reported for mood, fatigue, headaches, cognition/memory, sleep, and noise sensitivity between the concussion and control groups. Although vision morbidities were prevalent in both groups, they were not significantly different between the two. (Figure 2).

**Figure 2:**
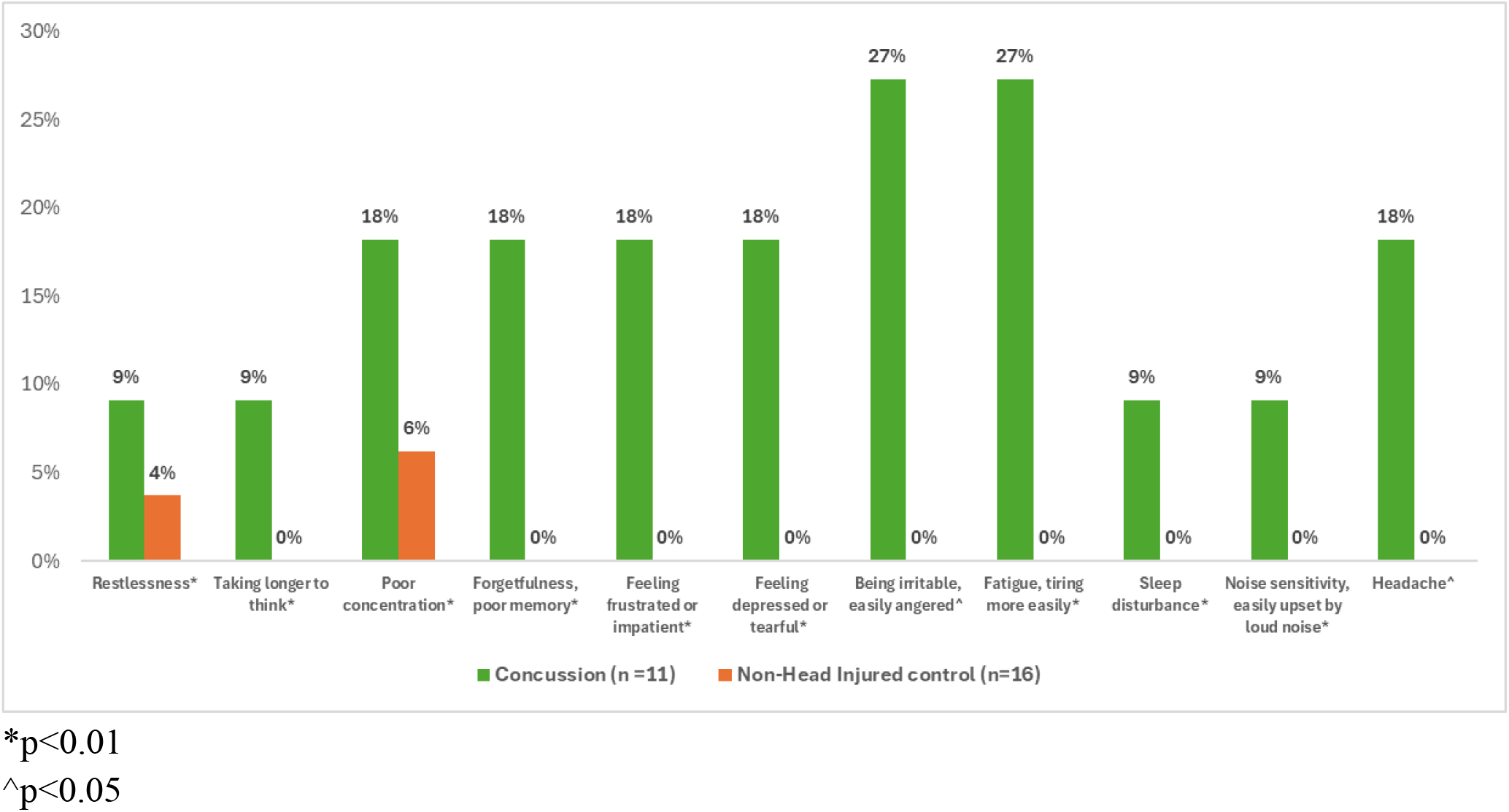
Domains of the RPQ with concussion patients endorsing “A severe problem” compared to non-head injury controls, 1-2 years (on average) post-concussion.

**Figure 3:**
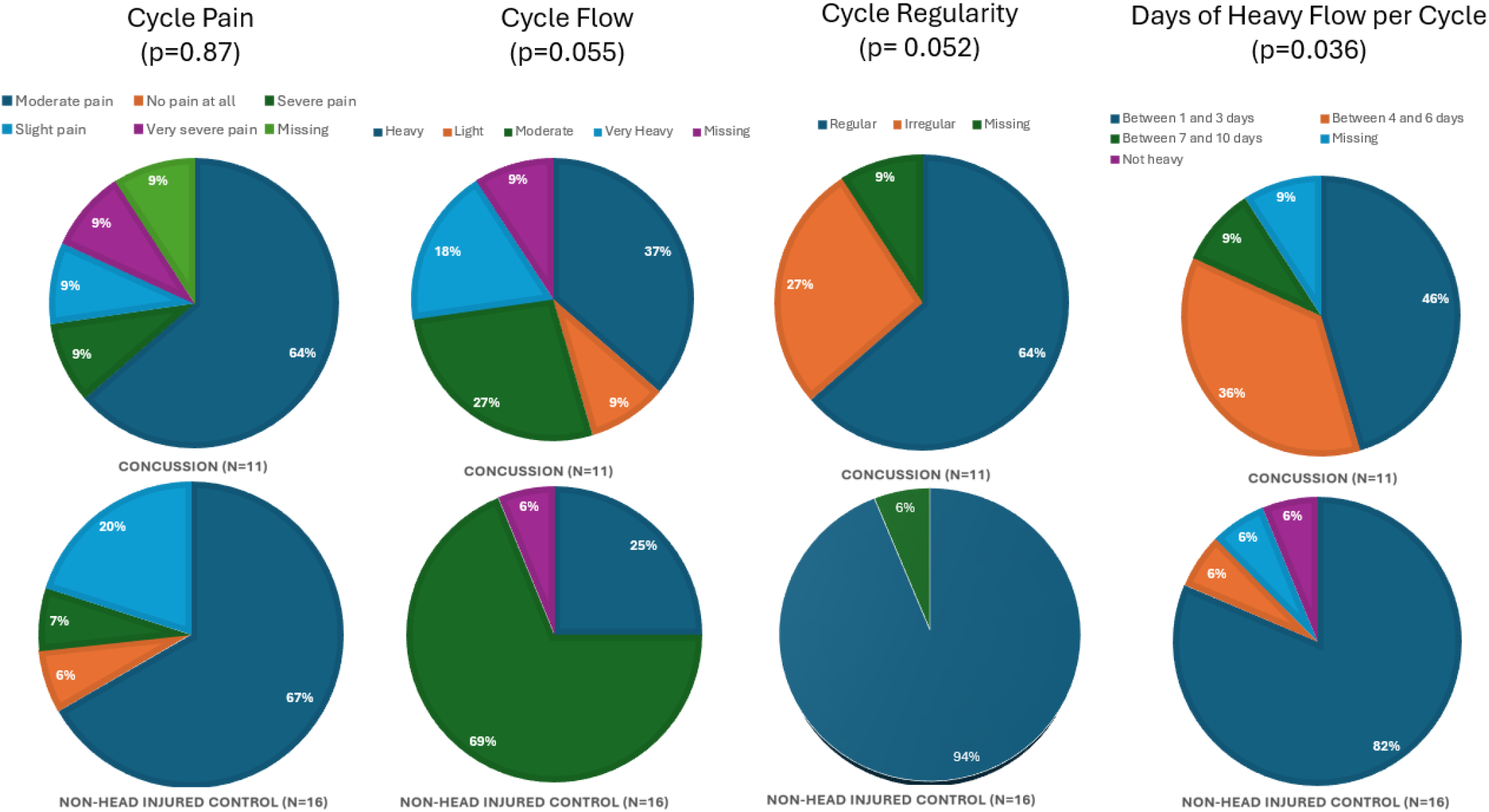
Menstrual cycle experiences of Concussion (n=11) and non-head injured controls (n=16), 1-2 years (on average) post-concussion.

**Figure 4:**
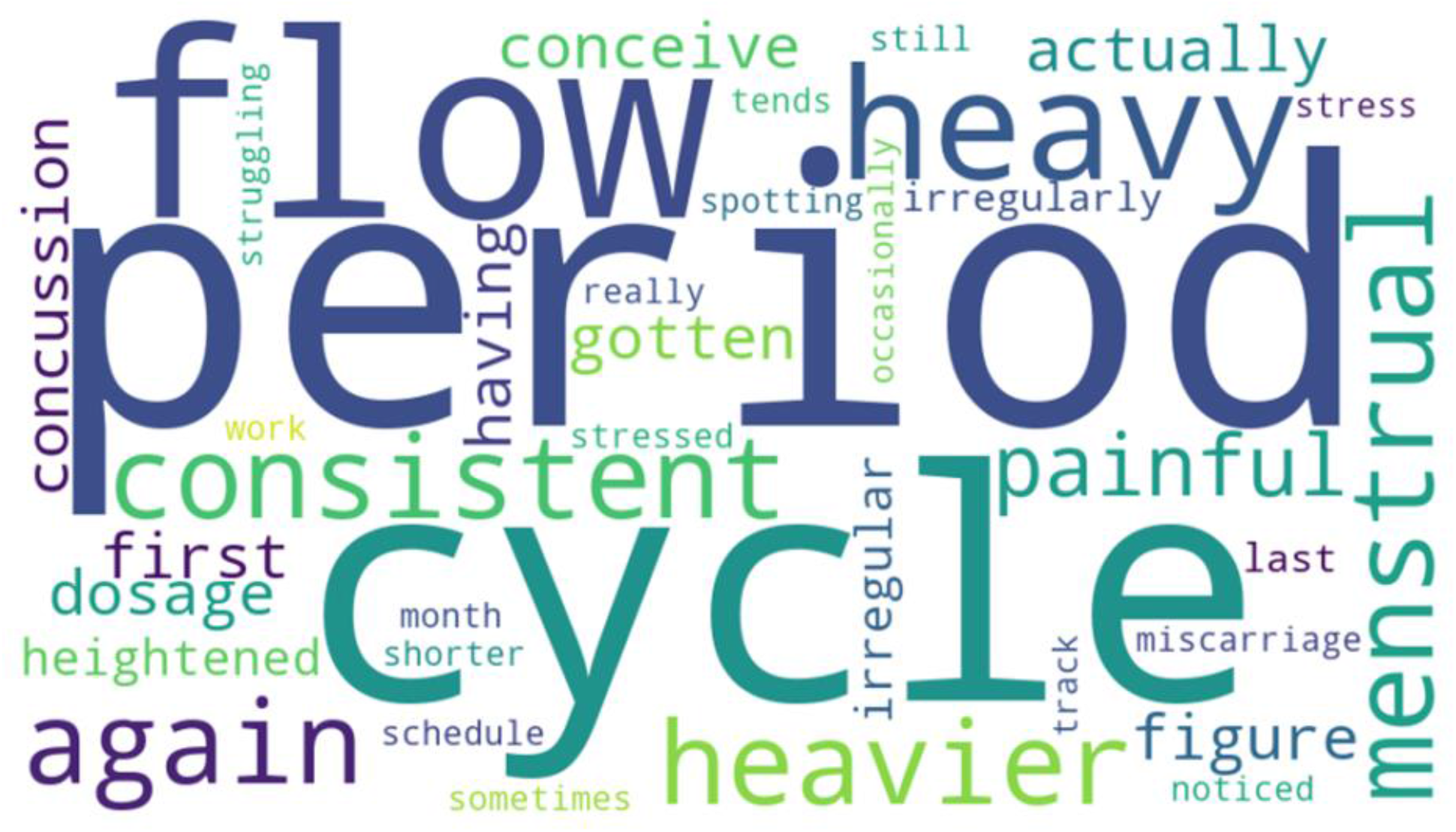
Word cloud visualization of emergent themes describing menstrual cycle experiences of concussion patients, 1-2 years (on average) post-concussion. Weight and size correspond to the number of mentions (n=7).

### Aim 2: Menstrual Patterns, Experiences, and Changes in Bleeding Severity

Compared to healthy controls, concussion patients reported significantly more days of heavy cycle flow. Whereas 82% of the controls reported between 1 and 3 heavy cycle days, only 46% of the concussion group had this experience; the majority had between 4-10 days of heavy menstrual bleeding each cycle; and none of them reported “not heavy” compared to 6% of the controls who did (p=0.0356). Of clinical relevance, more concussion patients reported severe/very severe cycle pain, very heavy flow, and more irregular cycles. A greater proportion of controls than concussion patients reported no cycle pain at all.

Qualitative responses and the word cloud visualization of participant narratives reinforced these findings. Comments related to “flow” “heavy” “heavier” and “painful” were predominant. In participants’ own words:

“Periods are more irregular and painful” (Response #6)

“Heavier and it lasts up to 10 days. Sometimes I get my period 2x in a month. “ (Response #9)

“Had a miscarriage and am now struggling to conceive again” (Response #5)

A few noted that there had been no cycle impacts, and one noted a trend towards recovery

“I was having periods irregularly due to stress and now they are on a very consistent schedule!”

(Response #7)

## Discussion

This study used a mixed-methods approach to examine the long-term impact of concussion on women’s lived experiences, with a particular focus on persistent symptoms, mental health, and menstrual and reproductive health. Compared with non-head injured controls, women with a history of concussion reported greater symptom burden, poorer mental health, and more menstrual health concerns, while qualitative findings demonstrated the lasting impact of these changes on daily life and overall quality of life.

While concussion has been commonly labeled as a transient injury, findings from this study suggest that the impact of concussion on women’s health and quality of life may persist long after the initial recovery stages.^38^ Quantitative analyses demonstrated significantly higher post-concussive symptom burden, depression, and anxiety among women with a history of concussion compared to non-head injured controls, while qualitative responses provided important context regarding how these symptoms continue to shape daily life long after the initial injury. Together, these findings suggest that concussion may have lasting and multifaceted consequences for women that extend well beyond the acute recovery period and highlight the importance of incorporating patient experiences alongside traditional clinical measures.

Consistent with prior literature, women with a history of concussion reported significantly greater post-concussive symptom burden and poorer mental health outcomes than controls.^14^ Participants frequently described ongoing challenges with headaches and migraines, visual impairments, concentration, and emotional regulation that continued to affect daily functioning years after injury. Although concussion is often characterized as a transient injury, our findings support growing evidence that a subset of individuals experience persistent symptoms that affect cognitive, emotional, and physical functioning long after the initial injury. Importantly, participants described these symptoms not as isolated clinical complaints but as factors that influenced nearly every aspect of their lives, including work, relationships, and recreation. The qualitative findings further demonstrated that concussion can alter how women perceive themselves and interact with the world around them. Participants described fear of engaging in activities they previously enjoyed, concerns regarding reinjury, and feelings of being fundamentally changed by their concussion experience. Statements such as “I’m scared to play hockey” and “I have a fear of stairs” reflect more than symptom persistence; they illustrate disruptions in their everyday routines and participation in meaningful activities. This loss of self may negatively affect psychological well-being and is consistent with the elevated rates of depression and psychological distress reported among individuals with persistent post-concussive symptoms.^39^ Recovery from concussion therefore appears to involve more than symptom resolution alone; it may also require individuals to adapt to changes in their confidence and sense of identity.

Importantly, our findings challenge assumptions that persistent symptoms following concussion in women are mostly due to *somatization*, or the tendency to experience and communicate psychological distress in the form of physical symptoms.^40^ Research from the chronic illness literature shows that women’s symptoms are more likely to be dismissed, minimized, or attributed to psychological causes compared to men,^41^ contributing to delays in diagnosis, inadequate treatment, and poorer health outcomes.^42^ Qualitative studies further show that women frequently feel unheard or invalidated within clinical encounters, particularly when symptoms are complex, subjective, or unexplained through existing diagnostic frameworks.^43^ These challenges may be particularly relevant in the context of concussion, where symptoms are often invisible and recovery trajectories highly individualized.^44,45^ As a result, persistent symptoms may be attributed to somatization, dismissed, and/or or undiagnosed. In contrast, participants in the present study described tangible and substantial disruptions across multiple domains of daily life, including cognition, physical function, work performance, social participation, and reproductive health that demonstrate the real-world consequences of persistent symptoms. Rather than reflecting perceived illness alone, these narratives suggest that ongoing difficulties following concussion represent real functional impairments that can affect overall well-being and quality of life that should be taken seriously; particularly for concussion patients who are women.

Beyond psychosocial and economic consequences, persistent symptoms may contribute to downstream physical health effects. Participants described avoiding sports, exercise, or other activities due to ongoing symptoms or fear of reinjury; such avoidance may have unintended consequences for long-term health. These findings have been reaffirmed in previous literature where patients report higher levels of exercise intolerance due to general deconditioning, post-injury psychological distress, and symptom exacerbation.^46,47,48,49^ Reduced participation in physical activity and increased exercise intolerance can independently increase risk for obesity, cardiovascular disease, and other chronic health conditions.^50,51,52^ This finding is particularly noteworthy given growing evidence that aerobic exercise is an effective treatment modality for persistent post-concussive symptoms and can improve symptom burden, exercise tolerance, and overall functioning and quality of life.^53,54,55^ By limiting engagement in physical activity, persistent symptoms may affect multiple aspects of physical health, supporting the view that concussion can have broader, whole-body consequences that persist longer than initial injury.

The effects of these persistent symptoms may also extend beyond individual health outcomes to broader socioeconomic consequences. Difficulties with concentration, memory, fatigue, and executive functioning are common persistent symptoms of concussion and have been associated with impaired academic achievement and reduced workplace productivity and performance.^11,56,57^ In concordance with this, participants described challenges maintaining focus and attention, affecting their ability to function at work. The economic burden of these lasting effects may be substantial; recent estimates suggest that productivity losses attributable to non-fatal traumatic brain injuries exceeded $19 billion in the United States in 2023.^58^ Although economic outcomes were not directly measured in this study, participants’ experiences illustrate the mechanisms through which persistent cognitive and psychological symptoms may contribute to these substantial costs. These findings further emphasize that the burden of concussion extends beyond clinical outcomes and can affect other pertinent parts of well-being, including educational attainment, career progression, and financial stability.

Another unique contribution of our study is the examination of reproductive and menstrual health experiences following concussion. Participants reported menstrual irregularities, changes in cycle characteristics, and concerns regarding fertility, reproductive health, and sexuality up to 2 years after their concussion. These findings align with growing evidence suggesting that concussions may influence neuroendocrine function through disruption of hypothalamic-pituitary pathways, which can worsen to produce significant long-term consequences.^18^ The sex-specific health concerns reported by participants highlight an often overlooked dimension of concussion recovery that may substantially affect the lived experiences of women. Given the importance of menstrual health as a marker of physical, mental, and social well-being, these findings underscore the broader implications of concussion for women’s health and suggest that reproductive health may warrant greater consideration within post-injury care.^59^ The apparent gap between these reported experiences and current concussion care raises important questions regarding how women’s health concerns are overall recognized and addressed within the healthcare system.

Historically, concussion research has focused predominantly on male populations, and many aspects of women’s health remain underrepresented in clinical guidelines and patient education materials.^60^ As a result, women may not receive counseling that adequately reflects the range of symptoms and concerns they experience following injury. Incorporating patient narratives into both research and clinical care may help bridge this gap by ensuring that outcomes important to women are recognized, validated, and addressed throughout recovery. A more holistic and patient-centered approach to concussion management is needed for female patients. Although the role of reproductive health counseling following concussion remains largely unexplored, the concerns regarding menstruation, fertility, and sexuality reported by participants suggest that this may represent an important yet underrecognized aspect of post-injury care for women. Discharge instructions and follow-up care may benefit from including education regarding potential mental health symptoms,^15,61,62^ menstrual cycle changes,^18^ sexual health concerns,^63^ and strategies to safely maintain physical activity during recovery.^64,65^ Recognizing and addressing the variety of experiences may help ensure that concussion care, as developed and practiced by researchers and clinicians, reflects not only the resolution of symptoms but also the broader realities of recovery as they fit into their patients’ lives. Additionally, blood-based, digital, and neuroimaging biomarkers offer promising opportunities to improve diagnosis, risk evaluation, and prediction of recovery trajectories.^66,67^ Given the heterogeneous nature of PCS symptoms and the continued reliance on self-reported symptoms measures in concussion assessment, such objective biomarkers may provide valuable complementary evidence of injury and recovery.^13,68^ Importantly, these measures should be developed alongside—not in place of—patient-reported and qualitative data to ensure that meaningful impacts on daily functioning, quality of life, and women’s health remain central to concussion care and research.

### Strengths & Limitations

A few limitations should be considered when interpreting these findings. Even though our mixed methods approach provided rich data, the relatively small sample size may introduce selection bias and limit generalizability, as individuals experiencing more severe or persistent symptoms may have been more likely to participate in the study. Additionally, participants were recruited from a single healthcare system and may not be representative of all women with concussion in the broader population. Further longitudinal research with larger and more diverse samples is needed to validate these findings and further investigate underlying neurobiological and psychosocial pathways.

Despite these limitations, our study also has several strengths. A key strength is the use of mixed-methods design, which integrates quantitative assessments of post-concussive symptom burden and mental health outcomes with qualitative data capturing participants’ lived experiences. This approach allowed for a more comprehensive understanding of concussion recovery, in a way that addresses more generalizable and comparative symptoms measures with opportunity to contextualize within the everyday scope of participant experiences through deeper explanation. The inclusion of qualitative responses was particularly valuable in identifying domains that may be underrepresented in standardized clinical assessments. Another strength is the focus on women’s experiences following concussion, a population that has historically been underrepresented in traumatic brain injury research. By centering female participants, this study highlights gender-specific aspects of recovery that are often overlooked in clinical guidelines and contributes to a more nuanced understanding of how concussion recovery overlaps with social determinants of health. Another strength is the longitudinal assessment of outcomes, which distinguishes this study from several others. By following up with women 1 to 2 years after their initial head injury, our findings contribute to evidence that concussions are neither “mild” nor transient. The injury can impact multiple body systems; ranging from vision, mood, cognition, and sleep to menstruation, sexuality, and pregnancy. Such longer-term assessments of outcomes are crucial for predicting and tracking patients’ recovery.

### Conclusion

A mixed methods approach that incorporates patients’ lived experiences is important when examining experiences of concussion pertaining to women, who are not only at greater risk for persistent symptoms but also navigate recovery within the context of complex and often competing social roles, including employment, caregiving, and household responsibilities.^69^ Evidence from broader chronic illness literature demonstrates that women frequently experience disruptions in work, leisure, and daily functioning, often requiring adaptation or abandonment of valued activities.^70,71,72^ In addition, post-concussion recovery may intersect with gender-specific aspects of health and bodily identity, including menstrual cycle changes,^73^ pregnancy-related concerns,^74^ and alterations in sexual health and intimacy^75^—topics that are rarely comprehensively captured in standardized outcome measures but may significantly shape experiences and perceptions of recovery. These different dimensions underscore the importance of understanding concussion recovery as both a socially and biologically embedded experience, impacting perceived identity as a whole.^76^

## Data Availability

Data is available upon reasonable request from the authors.

